# Re-emergence of respiratory syncytial virus following the COVID-19 pandemic in the United States: a modeling study

**DOI:** 10.1101/2021.07.19.21260817

**Authors:** Zhe Zheng, Virginia E. Pitzer, Eugene D. Shapiro, Louis J. Bont, Daniel M. Weinberger

**Author notes:** Corresponding author: Zhe Zheng, Department of Epidemiology of Microbial Diseases, Yale University, 60 College Street, New Haven, Connecticut, USA.

## Abstract

**Importance:** Respiratory syncytial virus (RSV) is a leading cause of hospitalizations in young children. RSV largely disappeared in 2020 due to precautions taken because of the COVID-19 pandemic. Projecting the timing and intensity of the re-emergence of RSV and the age groups affected is crucial for planning for the administration of prophylactic antibodies and anticipating hospital capacity.

**Objective:** To project the potential timing and intensity of re-emergent RSV epidemics in different age groups.

**Design, Setting, Participants:** Mathematical models were used to reproduce the annual RSV epidemics before the COVID-19 pandemic in New York and California. These models were modified to project the trajectory of RSV epidemics in 2020-2025 under different scenarios with varying stringency of mitigation measures for SARS-CoV-2: 1) constant low RSV transmission rate from March 2020 to March 2021; 2) an immediate decrease in RSV transmission in March 2020 followed by a gradual increase in transmission until April 2021; 3) a decrease in non-household contacts from April to July 2020. Simulations also evaluated factors likely to impact the re-emergence of RSV epidemics, including introduction of virus from out-of-state sources and decreased transplacentally-acquired immunity in infants.

**Main Outcomes and Measures:** The primary outcome of this study was defined as the predicted number of RSV hospitalizations each month in the entire population. Secondary outcomes included the age distribution of hospitalizations among children <5 years of age, incidence of any RSV infection, and incidence of RSV lower respiratory tract infection (LRI).

**Results:** In the 2021-2022 RSV season, we expect that the lifting of mitigation measures and build-up of susceptibility will lead to a larger-than-normal RSV outbreak. We predict an earlier-than-usual onset in the upcoming RSV season if there is substantial external introduction of RSV. Among children 1-4 years of age, the incidence of RSV infections could be twice that of a typical RSV season, with infants <6 months of age having the greatest seasonal increase in the incidence of both severe RSV LRIs and hospitalizations.

**Conclusions and Relevance:** Pediatric departments, including pediatric intensive care units, should be alert to large RSV outbreaks. Enhanced surveillance is required for both prophylaxis administration and hospital capacity management.

## Introduction

Respiratory syncytial virus (RSV) infection is a leading cause of acute respiratory hospitalizations in infants, young children, and the elderly.^1-3^ Individuals develop only partial immunity following RSV infections, and this incomplete immunity permits reinfections throughout life.^4^ Although most infants are born with protective immunity against RSV infections due to antibodies from the mother that are acquired transplacentally, this protection wanes quickly with time.^5-7^

RSV epidemics occur with notable spatiotemporal patterns in the United States, with consistent seasonal timing and duration.^8,9^ However, the incidence of RSV in many regions in the United States has declined since the introduction of mitigation measures for COVID-19 pandemic in March 2020.^10^ The low positivity rate continued throughout 2020 without an obvious increase in late fall and winter when seasonal RSV epidemics typically occur. Many other countries reported similarly low frequencies of RSV detection in the 2020 season.^11-13^

As mitigation measures have been gradually lifted, different patterns of RSV epidemics emerged in different regions in early 2021.^10-13^ Later-than-usual RSV activity was reported in France, Spain and in some U.S. states.^10,12,14^ Large out-of-season surges of RSV infections were reported in both Australia and South Africa.^15,16^ Nonetheless, even with many restrictions lifted, RSV activity remains low in many other countries.^17,18^ What causes these differences remains unclear.

The potential impacts of mitigation measures on the number, timing, and age of RSV hospitalizations are crucial for planning for the administration of RSV prophylaxis and for hospital utilization, since regular annual RSV epidemics often fully use hospital capacity in pediatric departments. However, many factors may affect when and to what extent RSV epidemics resume. For example, introduction of RSV from other regions may accelerate the process, and low immunity in the population due to a low incidence of reinfections may produce a large population of susceptible individuals sufficient to trigger an outbreak.

Our study aims to simulate a range of potential scenarios and to inform data collection for better epidemic projections of RSV activity in the U.S. In this study, we used historical RSV inpatient data, combined with validated transmission dynamic models, to evaluate the potential patterns of re-emergence of RSV epidemics with different stringencies of mitigation measures. We also examined how the age distribution of RSV infections and hospitalizations might be expected to change in the coming years following these different scenarios.

## Methods

### Data

RSV-specific hospitalization data for New York (2005-2014) and California (2003-2011) were obtained from State Inpatient Databases of the Healthcare Cost and Utilization Project maintained by the Agency for Healthcare Research and Quality.19 These comprehensive databases contain all hospital discharge records from community hospitals in participating states. Datasets include the month of hospitalization and the age of the patient. A hospitalization was defined as due to RSV if any of the discharge diagnostic codes included 079.6 (RSV), 466.11 (bronchiolitis due to RSV), or 480.1 (pneumonia due to RSV), based on the International Classification of Disease Ninth Revision [ICD-9].^20^ Information about population size in each age group was obtained from the US Census Bureau’s American Community Survey.^21^ Birth rate by year and state was obtained from CDC vital statistics.^22^ RSV surveillance data for California (2009-2018) were obtained from the Immunization Branch, California Department of Public Health and were used to validate model prediction.^23^ We also performed sensitivity analyses using parameters fitted to a similar inpatient dataset from Colorado to illustrate potential impacts of mitigation measures on biennial RSV epidemics.^24^

### Simulations

To predict RSV transmission dynamics, we extended a previously published RSV transmission model (see Supplement for details, Figures S1-S3).^24^ We generated forward simulations using varying input parameters (see Supplement for details), assuming the same immigration/emigration, death and birth rate as 2019. We used fixed control periods in our main analysis. We also considered various durations of the control periods and the impacts of mitigation measures on RSV transmission dynamics in states with different epidemic timing, including biennial epidemics, as sensitivity analyses.

We simulated the monthly number of RSV hospitalizations from 2021 to 2025. With many unknowns, our aim was to explore a wide range of possible scenarios (see Box 1) rather than making precise predictions.

#### Decrease in transmission

We evaluated a range of reductions in the RSV transmission rate, from 10% to 25%, based on the results of a previous study analyzing the impact of mitigation measures on seasonal respiratory viruses.^25^ We evaluated two kinds of decrease in transmission beginning in March 2020 and lasting until April 2021: 1) a constant decrease, and 2) a large decrease followed by a linear increase in transmission. The rationale for the second type of decrease comes from reports that mitigation measures were strictest at the beginning of the COVID-19 pandemic and were gradually relaxed with time.^26-28^

#### Changes in contact patterns

We also explored the impacts of heterogeneous changes in contact patterns on RSV epidemics. Specifically, we examined the effects of a three-month stay-at-home order from April 1 to July 1, 2020.^26,28^ We calculated the percentage of household contacts in detailed age groups and multiplied by the age-specific contact rates of respiratory-spread infectious agents in the corresponding age groups.^29-31^ We assumed an 82%^32^ decrease in non-household contacts and a 10%^33^ increase in household contacts.

#### Impact of virus introduction from external sources

Stay-at-home orders and travel restrictions may also have an effect on the introduction of RSV into a population. Travel restrictions between countries and states may limit the introduction of external RSV infections, while constant virus seeding from other regions may accelerate the re-emergence of RSV epidemics. There is little data on the rate of infections of RSV that are imported from other regions. Therefore, we explored a range of 1 to 10 imported infections per 100,000 population per month. We assumed these external infections are mild and do not lead to LRI or hospitalization. In the main analysis, we assumed the number of external infections decreased to 0 in April 2020 and gradually increased back to 40% of the original level in February 2021 based on changes in weekly total air travel.^34^ We also explored various other scenarios (see Supplement).^35^

#### Impact of decreased duration of transplacentally-acquired immunity in infants

Reduction in transmission results in low virus activity in the community, which may cause fewer exposure opportunities for the general population. Because virus exposure can boost immunity levels,^36^ pregnant women may have lower concentrations of antibodies because of lack of boosting from a lower rate of exposure. Correspondingly, the level and duration of the immunity in infants acquired from their mothers may also decrease. Since few previous studies quantified how virus exposure in pregnant women may affect the duration of transplacentally-acquired immunity in infants, we probed the effect of a lack of boosting by proposing a range of possible decreases in the duration of transplacentally-acquired immunity on top of the linear change in RSV transmission. The period of shortened transplacentally-acquired immunity is therefore dependent on the RSV epidemics, from November 2020 to October 2021 based on observations and the results of simulations.

### Outcome Measures

The primary clinical outcome of interest is the predicted monthly number of RSV hospitalizations. Secondary outcomes of interest included the age distribution of hospitalizations among children under 5 years of age, incidence of any RSV infection, and incidence of RSV lower respiratory infection. Percentage changes in incidence were calculated by comparing the differences between the predicted incidence in the 2021-2022 RSV season with and without changes to the transmission rate related to mitigation measures (see Supplements for details).

## Results

### Expected timing of the re-emergence of RSV under different scenarios

Under Scenario 1, we assumed that transmission decreased homogeneously by 20% from March 2020 to March 2021 without any external source of infections. In this case, the model predicted an absence of RSV epidemic in 2021 and an out-of-season outbreak starting in the summer of 2022. Models with constant low transmission took more than a year to resume annual epidemics (Figure 1A).

**Figure 1.**
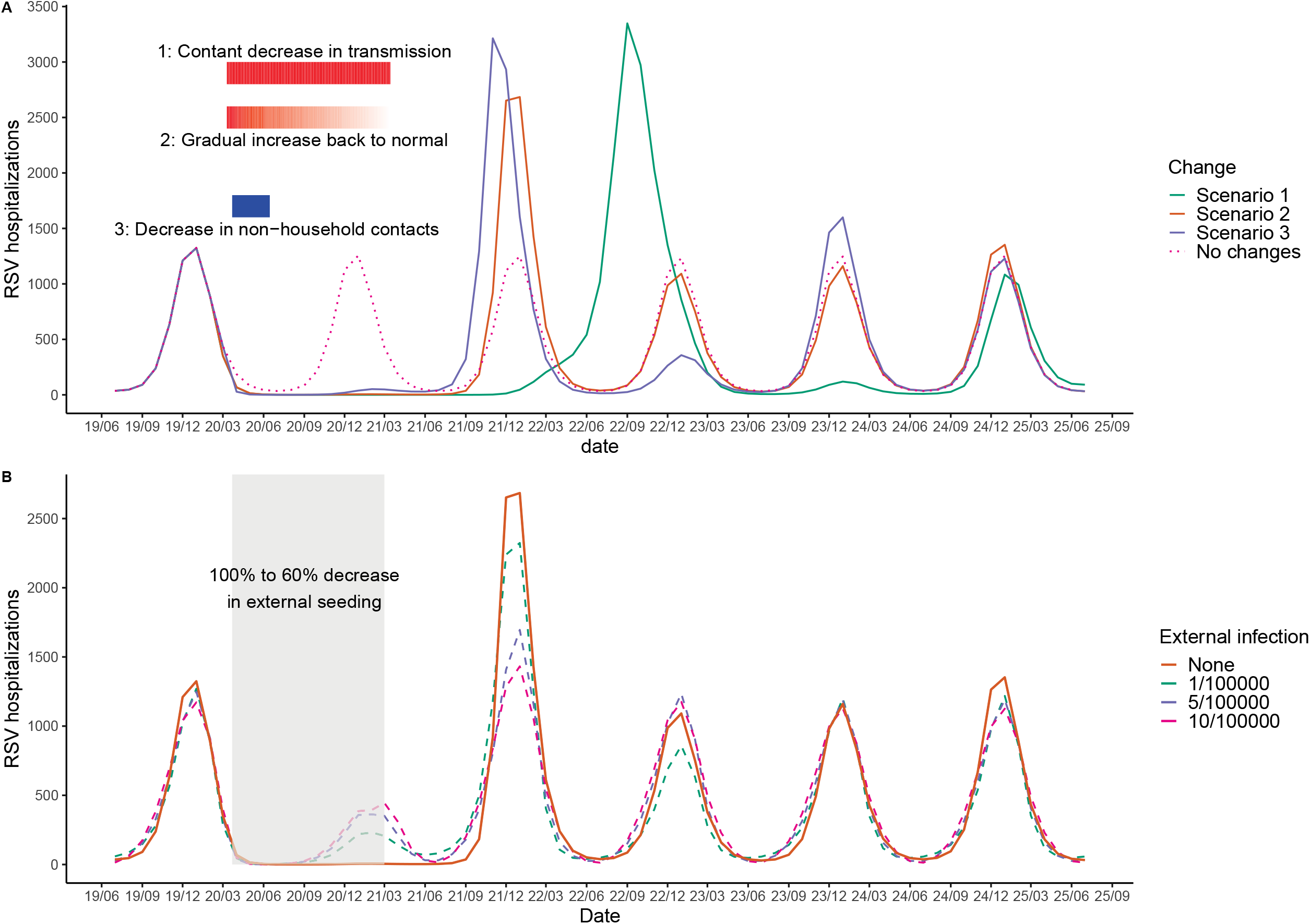
Expected RSV hospitalizations under different scenarios, New York, 2019–2025. A) The impacts of mitigation measures on RSV epidemics. The dotted dark pink line shows the counterfactual scenario that there is no COVID-19 pandemic and no mitigation measures in place. The solid lines show three scenarios of stringency of mitigation measures. The green line represents Scenario 1: 20% constant decrease in transmission from March 2020 to March 2021. The orange line represents Scenario 2: a sudden 20% decrease in RSV transmission in March 2020 followed by a linear increase back to normal. The purple line represents Scenario 3: 82% decreased non-household contacts and 10% increased household contacts between April and July 2020. The red rectangle on the top, the gradient red rectangle in the middle and the blue rectangle on the bottom indicate the length and the stringency of Scenarios 1-3, respectively. B) The impacts of importation of external infections on Scenario 2. The solid orange line is the same as the orange line in panel A), corresponding to Scenario 2. The dashed lines represent a range of background importations of external infections. The grey shaded area indicates a 100% to 60% decrease in external infections because of the reduced air traffic between March 2020 to March 2021.

Compared with the constant low transmission rate scenario, a sudden decrease in RSV transmission in March 2020 followed by a linear increase over a 13-month-period (Scenario 2) allowed for sufficient infections to trigger a large outbreak in the winter of 2021-2022. With this scenario, we would expect to see epidemics return to annual cycles in the winter of 2022-2023 (Figure 1A). Models with smaller decreases in transmission generally generated earlier re-emergence of RSV epidemics (Figure S4).

In the third scenario, a three-month stay-at-home order beginning in April 2020 substantially reduced the number of non-household contacts and resulted in a delayed, small uptick in RSV infections. With these conditions, infections re-emerge around February-March 2021, rather than the expected November 2020. The RSV epidemic in the 2021-22 season is expected to have an earlier onset and peak timing than the usual RSV season under this scenario (Figure 1A).

The scenarios described above assume no external introduction of RSV infections. In contrast, if Scenario 2 (20% linear change in transmission) was modified to allow for the external introduction of infections during the control period (Scenario 4), a small increase in RSV hospitalizations would have been expected in the winter and spring of 2020-2021. This increase is expected to be followed by a small spike corresponding to a return to normal virus importation due to lifted restrictions on inter-state travel. Based on this scenario, we expect an earlier onset, higher peak number and more patients per season in the 2021-2022 than a typical RSV season (Figure 1B).

Based on our simulation (Scenario 5), shorter transplacentally-acquired immunity in infants did not substantially change the timing or the amplitude of RSV epidemics on top of the changes resulting from reduced transmission (Figure S5).

### Shifts in the age distribution of infection during the re-emergence of RSV

As reported, mitigation measures were gradually relaxed in most of the states.^26-28^ Therefore, we chose Scenario 2 to illustrate changes in the age distribution of infection during the re-emergence of RSV. The following findings are consistent across scenarios although the exact numbers are different. Under Scenario 2, the average age of hospitalization among children under 5 years is expected to be higher in the 2021-2022 RSV season compared with preceding years. The average age of hospitalization is predicted to gradually return back to the pre-pandemic level in 2023 (Figure 2). Young children are expected to have higher incidence of hospitalizations this upcoming epidemic year (July 2021 to June 2022).

**Figure 2.**
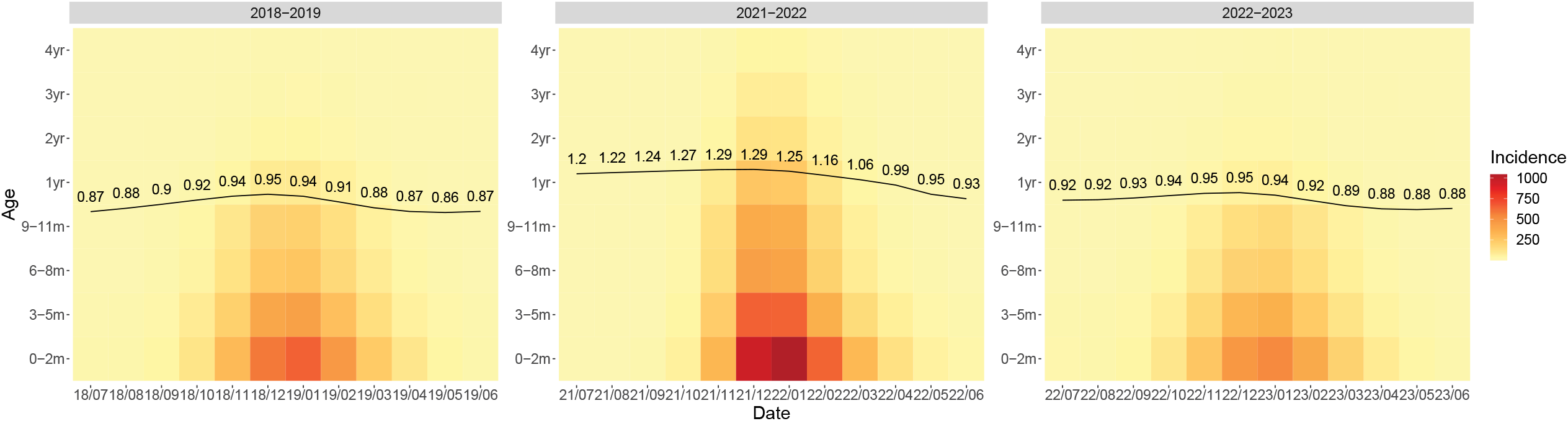
The average age of RSV hospitalization among children under 5. The background color represents the incidence of RSV hospitalization in each age group in each month. Darker red colors indicate a higher incidence. The black line and values indicate the average age of hospitalization varies with time.

Across all age groups, the model predicts an 87% increase in RSV LRIs and an 83% increase in RSV hospitalizations during the 2021-2022 epidemic year under Scenario 2 compared with a typical RSV season. Looking into the details of the age distribution for different clinical outcomes, children aged 1 to 4 years are expected to have the greatest percentage increase in the incidence of RSV infection, lower respiratory infection (LRI) and hospitalization compared with a typical pre-pandemic RSV season (Figure 3). However, infants age 3 to 5 months old are expected to have the largest incidence of LRI, and infants under 3 months are expected to have the largest incidence of RSV hospitalization.

**Figure 3.**
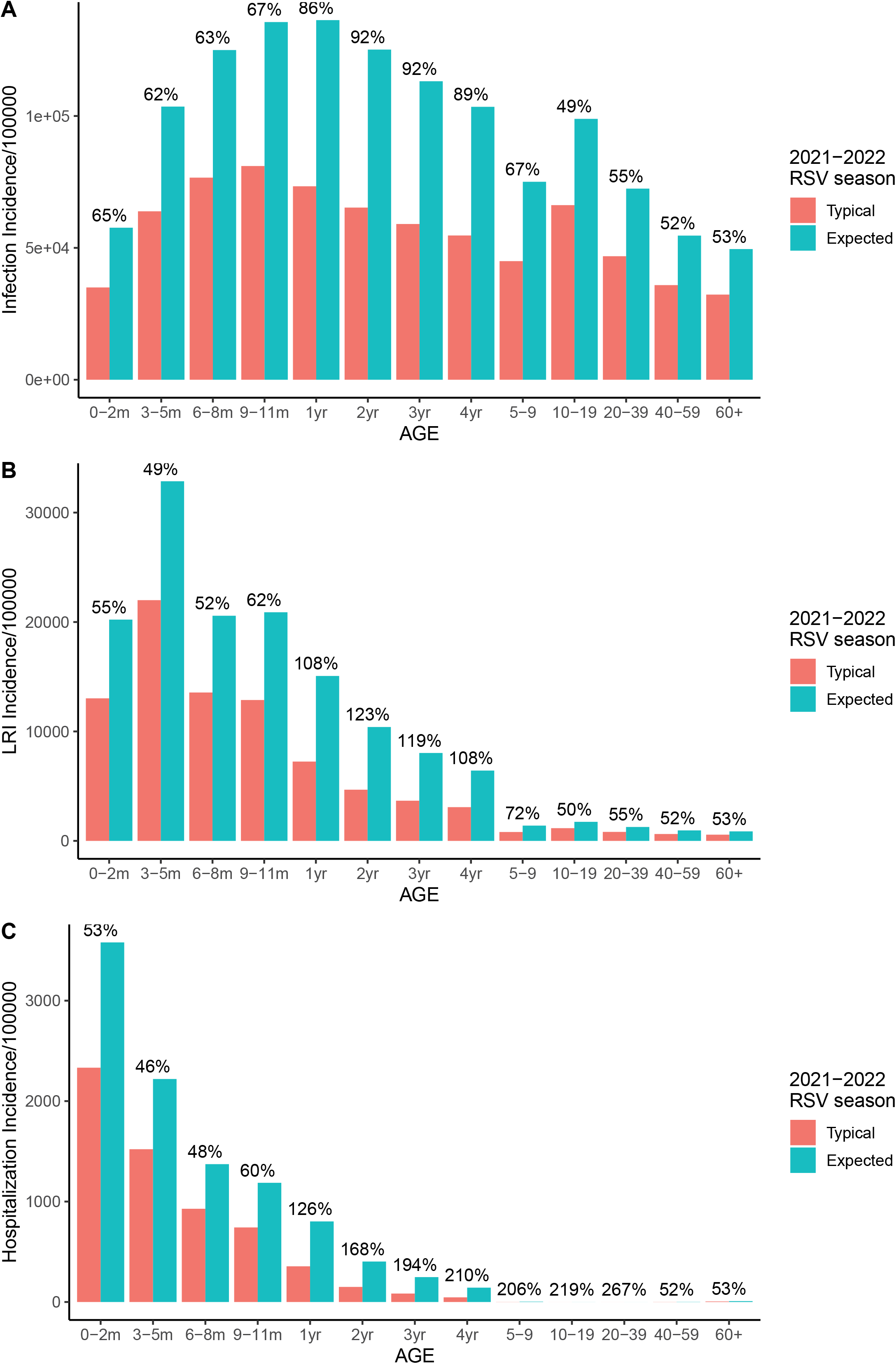
Age distribution of RSV infections, LRIs and hospitalizations, 2021-2022 RSV season. Panels A to C correspond to RSV infections, RSV LRIs and RSV hospitalizations, respectively. The red bars show the counterfactual incidence of RSV cases during the 2021-2022 RSV season if there was no COVID-19 pandemic and no mitigation measures in place. The blue bars show the expected incidence of RSV cases under Scenario 2 during the 2021-2022 RSV season. The numbers on the top show the percentage difference between the expected incidence and the counterfactual incidence in each age group.

## Discussion

Seasonal RSV epidemics have been interrupted by COVID-19 mitigation measures during 2020-2021. Understanding the potential timing, intensity and age distribution of re-emergent RSV epidemics is crucial for clinical and public health decision-making. Our results suggest that stricter mitigation measures will lead to later and larger epidemic outbreaks and a shift in the age distribution due to an accumulation of susceptibility within the population. However, the importation of infections into a population may accelerate the re-emergence process and cause an earlier-than-usual outbreak in the upcoming RSV season.

Variation in the patterns of re-emergent RSV epidemics across regions and countries could potentially be explained with our simulated results. (1) In Australia, an island country, closing the borders and implementing a mandatory quarantine were probably sufficient to eliminate imported infections. Australia also implemented a one-month stay-at-home order in the beginning of the COVID-19 pandemic and gradually relaxed all other restrictions within three months. These short-term mitigation measures may have temporarily suppressed local RSV transmission, and the accumulation of susceptibility since the end of the previous RSV season in October 2019 may have triggered the large out-of-season outbreak. Our simulated average age of hospitalizations is similar to the reported median patient age in Australia both before the pandemic and during the re-emergent RSV epidemic.^37^ (2) In New York, which has many and frequent connections to the other parts of the U.S. and the world, the risk of virus importation likely existed during the COVID-19 travel restriction period. The risk of virus importation depends both on the volume of travel and the level of external RSV activity. Therefore, the increase in virus importation during the control period may not be linear as travel volume increased. Before November 2020, most places in the world reported no RSV epidemics. Virus importation from other areas was therefore highly unlikely. This may be the reason that we observed a later and steeper increase in RSV epidemics during the spring of 2021 (see Figure S6). (3) In Argentina and Canada, governments implemented very strict mitigation measures at the start of COVID-19 pandemic.^38,39^ Some restrictions have been extended and remained in place as of April 2021 to protect against new variants.^40,41^ It is worth noting that these two countries represent both the northern and southern hemisphere and have opposite RSV seasons.^42^ Nonetheless, both countries reported low RSV activity as of April 2021.

In all of our scenarios, the upcoming RSV season is expected to have a higher-than-normal intensity. Our analysis suggests that infants <6 months of age are expected to have about 1.5 times increased risk of LRI and hospitalization as a result of RSV infections during the upcoming RSV season. At the same time, RSV infections in children aged 1 to 4 years are expected to double regardless of severity compared with a typical RSV season. This is alarming because pediatric facilities and intensive care units may be overloaded with patients in the upcoming RSV season.^43,44^ Even worse, the timing of re-emergent RSV epidemics varied among scenarios, making it difficult to predict the timing. Aberrations in timing may decrease the impact of palivizumab, since the administration of antibody prophylaxis for high-risk infants needs to be timed to coincide with the epidemic.^45^ Thus, enhanced, year-round surveillance for RSV infections is needed to inform the use of palivizumab over the 2021-2022 season. It is important for pediatric facilities and intensive health care units to be aware and prepared for these scenarios, especially since future RSV epidemics may occur at an unusual time.

Our study tested additional assumptions made by a previous study of the impact of mitigation measures on RSV dynamics by exploring a variety of scenarios, as well as considering the age distribution of cases.^25^ Our overall projections for upcoming RSV epidemics are similar: large outbreaks are very likely once mitigation measures are relaxed.

Our research suggests that virus introduction affects the timing and intensity of re-emergent RSV epidemics. Research from the late 1990s in the U.S. suggested that external sources of RSV infections may exist but are uncommon,^46,47^ in contrast to a recent finding from Kenya suggesting that RSV was reintroduced into communities every year from other parts of the country.^48^ With increased global connectivity, the mixing patterns of RSV transmission between and within communities in the U.S. may have changed. High-density sampling and sequencing of RSV across a region will be critical to understand the current level of importation of RSV infections.

For the re-emergent epidemics (2021-2022), our model (under Scenario 2 assumption) suggests an older average age of hospitalizations. This makes intuitive sense, since many children born in 2020 were spared from RSV infection due to the low virus activity; these children will be older when they get infected for the first time during the re-emergent epidemics. Consequently, the American Academy of Pediatrics should consider modifying prophylaxis guidelines to include high-risk infants <2 years of age for the 2021-2022 season. The average age of hospitalization will gradually decrease in the following RSV seasons because of the depletion of older susceptible individuals during the first re-emergent outbreak and the constant replenishment of susceptible infants through new births and waning transplacentally-acquired immunity.^49^

There are several caveats to our results. First, we did not have explicit data on the level of virus introduction or the effects of lack of boosting on the duration of protection provided to infants by transplacentally-acquired antibodies to RSV. Although we explored a range of values, it is possible that the real values are outside of the range in our models. Our model predictions are most sensitive to the level of RSV introduction. Additional data on these factors will be helpful for future modeling of RSV transmission dynamics. Research on the relationship between virus exposure and the duration of transplacentally-acquired immunity in infants may also help to explain discrepancies in the efficacy of maternal vaccines across different transmission settings.^50^ Second, we used historical inpatient data to fit our transmission models. However, the intensity and seasonality of RSV epidemics may have changed over the past few years. To address this possibility, we explored a variety of intensity levels and both annual and biennial cycles with data from different states in our sensitivity analysis (see Figure S7-S16). Also, ongoing RSV surveillance data from California,^51^ Florida,^52^ Minnesota,^53^ Oregon,^54^ and Texas^55^ suggests that RSV activity has been very consistent over recent years. Finally, there are other possible factors influencing re-emergent RSV epidemics, such as the impact of introduction of RSV vaccines and monoclonal antibody.^56^ As the extended half-life antibody against RSV, Nirsevimab, has been proved to be efficacious in phase III clinical trials,^57,58^ research on its impact may be important to elucidate future epidemics of RSV infections.

In conclusion, re-emergent RSV epidemics in 2021-2022 are expected to be more intense and to affect patients in a broader age range than in typical RSV seasons. The timing of re-emergent RSV epidemics may be different from the usual RSV season, depending on the duration of mitigation measures and the extent of virus introduction from other regions. Clinicians should be alert to the possibility of out-of-season RSV outbreaks.

## Data Availability

The demographic that support the findings of this study are publicly available from the American Community Survey of the U.S. Census Bureau. The hospitalization data are not available publicly but can be obtained upon signing a data use agreement with the Agency for Healthcare Research and Quality.RSV surveillance data for California (2009-2018) can be obtained from the Immunization Branch, California Department of Public Health.

## Other declarations

I confirm all relevant ethical guidelines have been followed, and any necessary IRB and/or ethics committee approvals have been obtained.

## Tables

### Box 1

**Description of the Five Simulations**

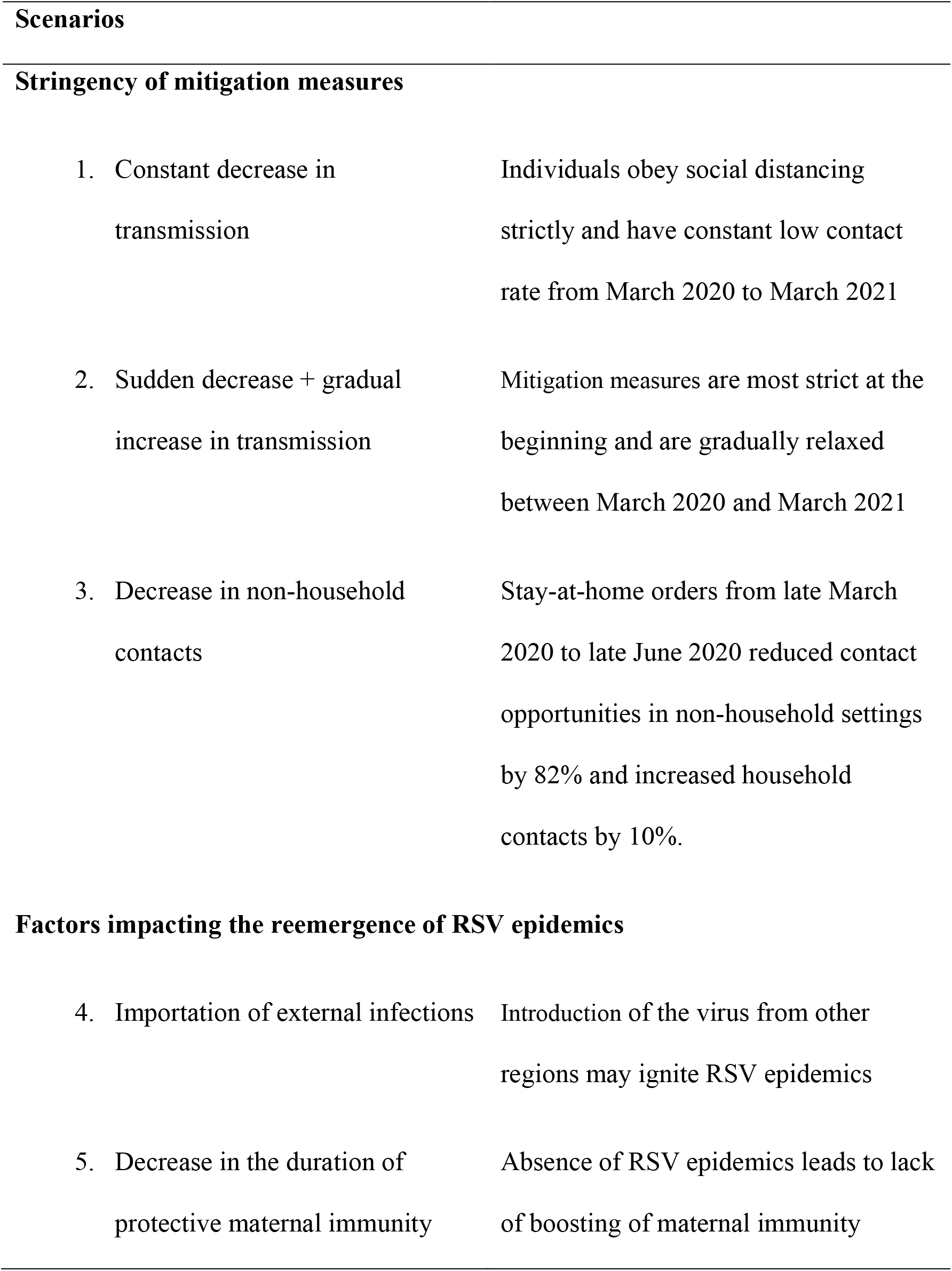

|  |  |
| --- | --- |
| 1. Constant decrease in transmission | Individuals obey social distancing strictly and have constant low contact rate from March 2020 to March 2021 |
| 2. Sudden decrease + gradual increase in transmission | Mitigation measures are most strict at the beginning and are gradually relaxed between March 2020 and March 2021 |
| 3. Decrease in non-household contacts | Stay-at-home orders from late March 2020 to late June 2020 reduced contact opportunities in non-household settings by 82% and increased household contacts by 10%. |
| 4. Importation of external infections | Introduction of the virus from other regions may ignite RSV epidemics |
| 5. Decrease in the duration of protective maternal immunity | Absence of RSV epidemics leads to lack of boosting of maternal immunity |

