## Supplementary document for "Re-emergence of respiratory syncytial virus following the COVID-19 pandemic in the United States: a modeling study"

### Supplementary Materials and Methods

#### Transmission dynamic models

Mathematical models were used to reproduce the annual RSV epidemics before the COVID-19 pandemic based on the inpatient data of New York (2005-2014) and California (2003-2011). Parameters to produce biennial RSV epidemics were taken from models fit to similar datasets from Colorado (1989–2009). This model assumes infants are born with transplacentally-acquired antibodies against RSV infections from their mothers (M). As transplacentally-acquired protective antibodies wanes, infants become susceptible to infection ($S_0$). Following each infection ($I_i$), individuals gain partial immunity that lowers both their susceptibility to subsequent infections and the duration and infectiousness of subsequent infections (see Figure S1). The force of infection for a specific age group $a$, $\lambda_a(t)$, for time $t$ is defined as:

$$\lambda_a(t) = \left(1 + b_1 \cos\left(\frac{2\pi t - \phi}{12}\right)\right) \sum_k \beta_{a,k} (I_{1,k}(t) + \rho_1 I_{2,k}(t) + \rho_2 I_{3,k}(t) + \rho_2 I_{4,k}(t))/N_k(t)$$

Seasonality in the force of infection is represented by $(1 + b_1 \cos(\frac{2\pi t-\phi}{12}))$, where $b_1$ is the amplitude of seasonality and $\phi$ is the seasonal offset. The chance of susceptible individuals in age group $a$ being infected is influenced by their contacts with infectious individuals in the entire population. $\beta_{a,k}$ is the transmission rate from age group $k$ to age group $a$. The proportion of infected individuals and their relative infectiousness at time $t$ is denoted by $(I_{1,k}(t) + \rho_1 I_{2,k}(t) + \rho_2 I_{3,k}(t) + \rho_2 I_{4,k}(t))/N_k(t)$, where $I_{1,k}$ is the number of infectious individuals of age $k$ during their first infection; $I_{2,k}$, $I_{3,k}$ and $I_{4,k}$ are the number of infectious individuals who have been infected two, three and four or more times, respectively; $\rho_1$ and $\rho_2$ denote the relative infectiousness of the second and subsequent infections; and $N_k$ is the total population of age $k$.

The transmission parameter $\beta_{a,k}$ can be further decomposed into the age-specific contact probability between age group $a$ and $k$ per unit time ($C_{a,k}$) and the probability of transmission given contact between an infectious and a susceptible individual ($q$). Age-specific mixing patterns were obtained from several previous studies, including detailed contact patterns for infants under 1 year of age and location-specific contact patterns.[1-3] Age was stratified into thirteen groups: infants younger than 3 months, 3-5 months, 6-8 months, 9-11 months, 1 year, 2 years, 3 years, 4 years, 5-9 years, 10-19 years, 20-39 years, 40-59 years, and ≥60 years.

The disease transmission process is linked to observation-level information. The probabilities of developing lower respiratory tract disease and being hospitalized upon RSV infection are informed by cohort studies conducted in the US and Kenya.[1,4-14] The number of lower respiratory tract infections (LRI) due to RSV is given by:

$$D_a(t) = \lambda_a(t)(S_{0,a}(t)d_{1,a} + \sigma_1 S_{1,a}(t)d_{2,a} + \sigma_2 S_{2,a}(t)d_{3,a} + \sigma_3 S_{3,a}(t)d_{3,a})$$

while the number of hospitalizations is given by:

$$H_a(t) = \lambda_a(t)(S_{0,a}(t)h_{1,a} + \sigma_1 S_{1,a}(t)h_{2,a} + \sigma_2 S_{2,a}(t)h_{3,a} + \sigma_3 S_{3,a}(t)h_{3,a})$$

where $\lambda_a(t)$ is the force of infection for a specific age group $a$ at time $t$ (as defined above). $S_{0,a}$ is the number of fully susceptible individuals of age $a$; $S_{1,a}$, $S_{2,a}$ and $S_{3,a}$ are the number of susceptible individuals who have been infected once, twice and more times, respectively. $\sigma_1$, $\sigma_2$ and $\sigma_3$ denote the relative risk of infection following the first, second, and more infections. $h_{1,a}$, $h_{2,a}$ and $h_{3,a}$ are the proportion of the first, second, and more infections that are hospitalized.

The average age of hospitalization among children under 5 in month $t$ is given by:[15]

$$A(t) = \frac{\sum P_a \lambda_a(t)(S_{0,a}(t)h_{1,a} + \sigma_1 S_{1,a}(t)h_{2,a} + \sigma_2 S_{2,a}(t)h_{3,a} + \sigma_3 S_{3,a}(t)h_{3,a})}{\sum \lambda_a(t)(S_{0,a}(t)h_{1,a} + \sigma_1 S_{1,a}(t)h_{2,a} + \sigma_2 S_{2,a}(t)h_{3,a} + \sigma_3 S_{3,a}(t)h_{3,a})}$$

where the weight $P_a$ is the midpoint of age group $a$.

Several model parameters were fixed based on data from previous cohort and modeling studies.[1,4-14] We used Bayesian inference to estimate the average duration of transplacentally-acquired immunity, age-specific probability of hospitalization in the 40-59 year and >60 year age groups, the transmissibility coefficient, and seasonal parameters by fitting the model to the hospitalization data from New York and California.[16,17] We identified the best-fit parameter sets by maximum a posteriori estimation.[18] The likelihood was calculated by assuming the observed number of hospitalizations in the entire population was Poisson-distributed with a mean equal to the model-predicted number of hospitalization, and that the observed age distribution was multinomial-distributed with probabilities equal to the model-predicted distribution of RSV hospitalizations in each age group.

To validate our model predictions, we fitted the transmission model to the inpatient data for California from 2003 to 2011; we then compared the model predictions with data on the percent of clinical specimens positive for RSV from a separate sentinel surveillance database from 2012 to 2018. We rescaled the percent positive data by calculating a scaling factor based on overlaying the surveillance data and inpatient data from 2009 to 2011 (see Figure S3).

We initialized the transmission models with 1 infectious individual in each age group (except for infants under 6 months) in July 1981 and used a burn-in period of 24 years and 22 years in New York and California, respectively. We also performed a sensitivity analysis around what re-emergence might look like in a state with a biennial pattern of epidemics, using parameters fitted to earlier data from Colorado as an example and assuming a linearly declining birth rate (from 17 to 10 births per 1,000 people per year). We used the same number of infectious individuals to initialize transmission model, and a burn-in period of 40 or 41 years starting from 1971 or 1970 to allow for greater incidence in even or odd years.

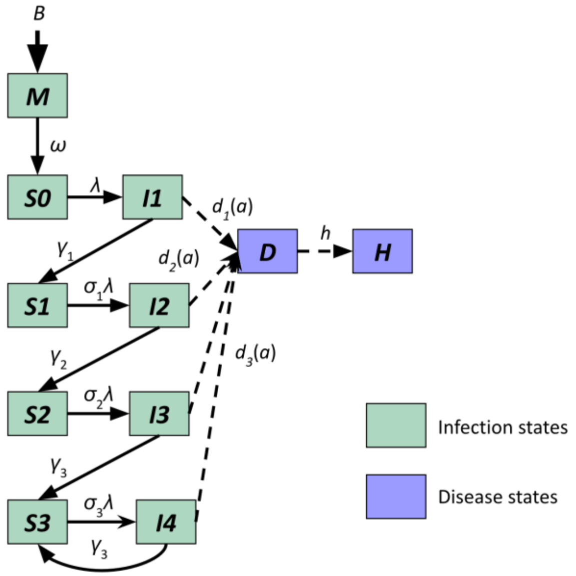


**Figure S1. Transmission dynamic model for RSV.** The green boxes represent infection states in the model, while purple boxes represent diseased states (RSV lower respiratory illness, D, and RSV hospitalizations, H).

#### Table S1 Shared transmission dynamic model parameters.

| Parameter description | Symbol | Parameter value | Reference for fixed or prior value | Note |
|---|---|---|---|---|
| Duration of transplacentally-acquired antibodies against RSV infections in infants from their mothers | $1/\Omega$ | 112 days | [19] | Fitted for NY and CA using maximum a posteriori estimation, assuming a Gamma(10,11) prior distribution |
| Duration of infectiousness | | | [20] | |
| First infection | $1/\gamma_1$ | 10 days | | |
| Second infection | $1/\gamma_2$ | 7 days | | |
| Subsequent infection | $1/\gamma_3$ | 5 days | | |
| Relative risk of infection following | | | [4,8,9,21] | |
| First infection | $\sigma_1$ | 0.76 | | |
| Second infection | $\sigma_2$ | 0.6 | | |
| Subsequent infection | $\sigma_3$ | 0.4 | | |
| Relative infectiousness | | | | |
| Second infections | $\rho_1$ | 0.75 | [4,8,10] | |
| Subsequent infections | $\rho_2$ | 0.51 | [22] | |
| Proportion of RSV infections leading to lower respiratory tract infection | | | [5] $\Pr(LRI\|I_s)$ <br> [23] $\Pr(I_s\|I)$ | The probability of lower respiratory infection (LRI) given infection was estimated as the product of LRI given symptomatic infection ($I_S$) times the probability of symptoms given infection: $\Pr(LRI\|I) = \Pr(LRI\|I_s) * \Pr(I_s\|I)$ |
| First infection, | | | | |
| 0-2 months old | $d_{p,0-2}$ | 0.44*0.9 | | |
| 3-5 months old | $d_{p,3-5}$ | 0.43*0.9 | | |
| 6-8 months old | $d_{p,6-8}$ | 0.23*0.9 | | |
| 9-11 months old | $d_{p,9-11}$ | 0.22*0.9 | | |
| 1-2 years old | $d_{p,1}$ | 0.21*0.8 | | |
| 2-4 years old | $d_{p,2}$ | 0.2*0.8 | | |
| ≥ 5 years old | $d_{p,adults}$ | 0.05 | [24] | |
| Second infection | $d_{s,a}$ | 0.5*$d_{p,a}$ | [8] | |
| Third+ infection | $d_{t,a}$ | 0.7*$d_{s,a}$ | [8] | |
| Proportion of RSV infections leading to hospitalization | | | [10,25,26] | |
| First infection, | $h_{p,0-2}$ | 0.20*$d_{p,0-2}$ | [27,28] | |

| | | | | |
|---|---|---|---|---|
| <3 months old | | | | |
| 3-5 months old | $h_{p,3-5}$ | 0.08*$d_{p,3-5}$ | | |
| 6-8 months old | $h_{p,6-8}$ | 0.07*$d_{p,6-8}$ | | |
| 9-11 months old | $h_{p,9-11}$ | 0.06*$d_{p,9-11}$ | | |
| 1-2 years old | $h_{p,1}$ | 0.06*$d_{p,1}$ | | |
| 2-4 years old | $h_{p,2}$ | 0.05*$d_{p,2-4}$ | | |
| $\geq 5$ years old | $h_{p,adults}$ | 0.02*$d_{p,adults}$ | | |
| Second infection | $h_{s,a}$ | 0.4*$H_{p,a}$ | [8] | |
| Third infection | $h_{t,a}$ | 0 except for the elderly | | Fitted for the elderly using maximum a posteriori estimation, assuming priors follow a uniform distribution U(0,1) |
| Scenarios for the impact of mitigation measures | | | | |
| Reduction in RSV transmission | | 10%-25% | [29] | |
| Decrease in non-household contacts | | 82% | [30] | |
| Increase in household contacts | | 10% | [31] | |
| External seeding during control period | | 100%-40% | [32] | |

#### Table S2 State-specific estimated transmission dynamic model parameters.

| | New York | California | Colorado |
|---|---|---|---|
| **Duration of maternal immunity** | 116.05 | 76 | 112 |
| **Basic reproductive number*** | 9.00 | 8.88 | 8.91 |
| **Amplitude of seasonality** | 0.16 | 0.25 | 0.24 |
| **Timing of seasonality** | 0.54 | 0.44 | 0.49 |
| **Reporting fraction** | 1 | 0.60 | 1 |

*The basic reproductive number ($R_0$) was estimated from $R_0 = \frac{\det(\beta_{a,k})}{\gamma_1} = \frac{\det(qC_{a,k})}{\gamma_1}$, using the next-generation matrix method; the parameter *q* was fitted to the data.

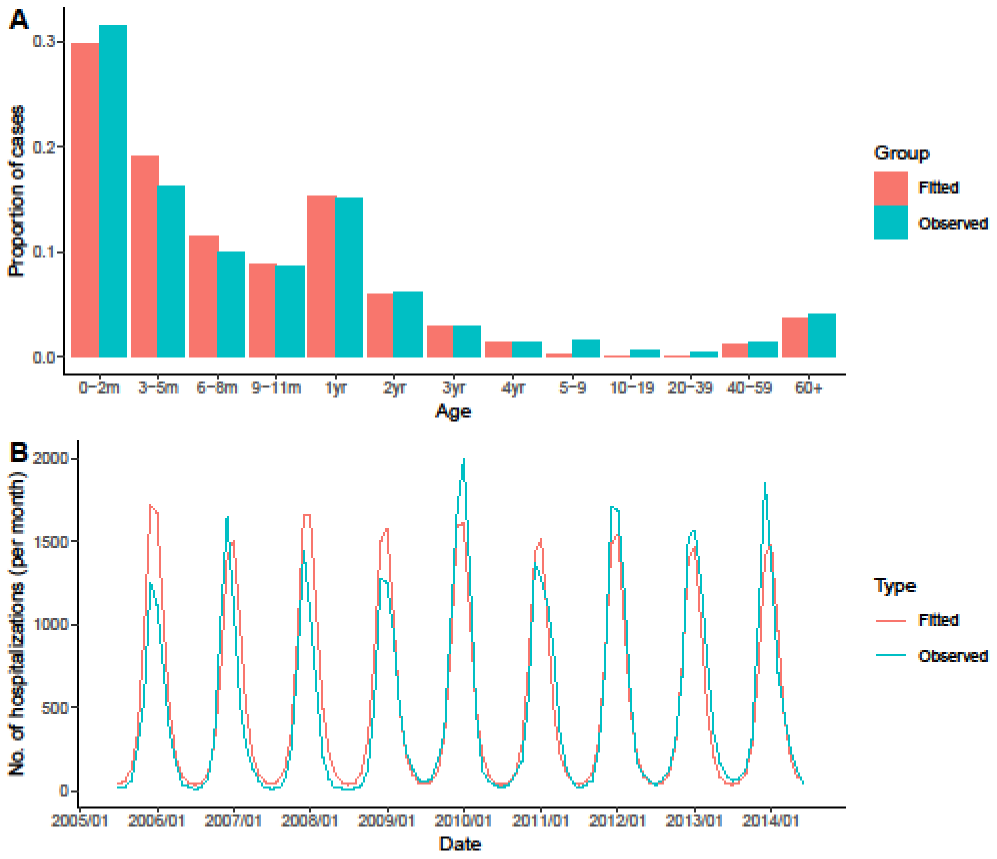


**Figure S2 Model fit to monthly RSV hospitalization data for New York.** The ICD9-CM coded hospitalization data is shown in blue and the fitted models are shown in red.

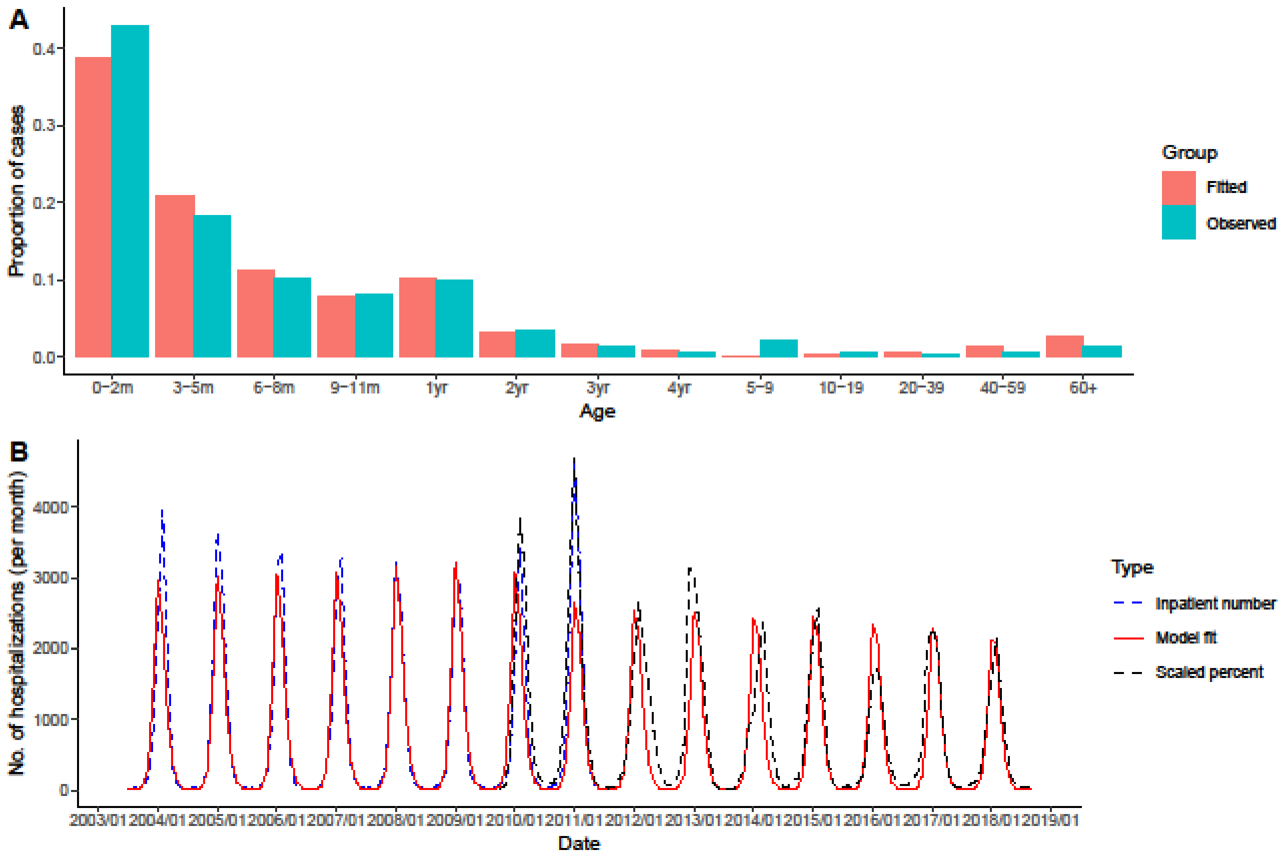


**Figure S3 Model fit to monthly RSV hospitalization data for California.** The ICD9-CM coded hospitalization data is shown in blue, the rescaled RSV positive percent data is shown in black, and the fitted models are shown in red.

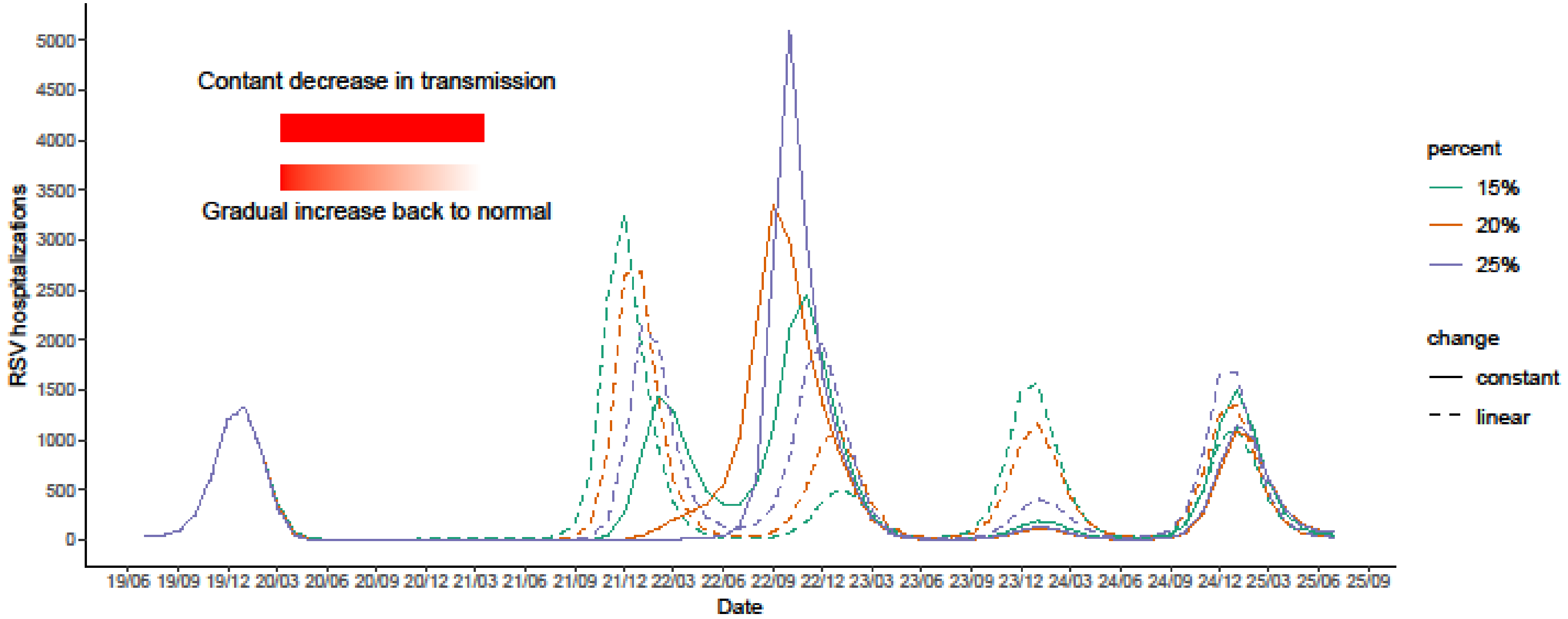


**Figure S4. Expected RSV hospitalizations under different stringency of mitigation measures.** The color lines indicate the percentage decrease under either constant decrease or linear change scenario. The solid lines represent a constant decrease in transmission. The dash lines represent the RSV hospitalization incidence under the assumption that mitigation measures are most strict at the beginning and are gradually relaxed. The solid red rectangle and the gradient red rectangle indicate the length of the change period is from March 2020 to March 2021.

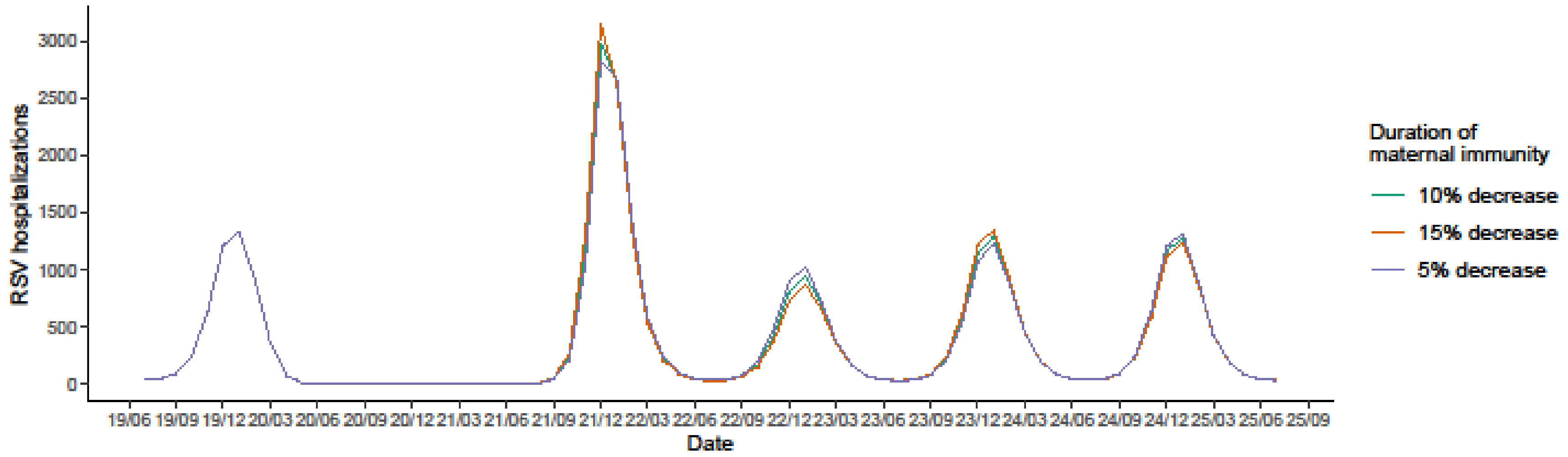


**Figure S5. The duration of transplacentally-acquired immunity in infants on RSV epidemics.** The colors of the lines show different percentage decrease in the duration of transplacentally-acquired immunity in infants as a result of lack of boosting in pregnant women.

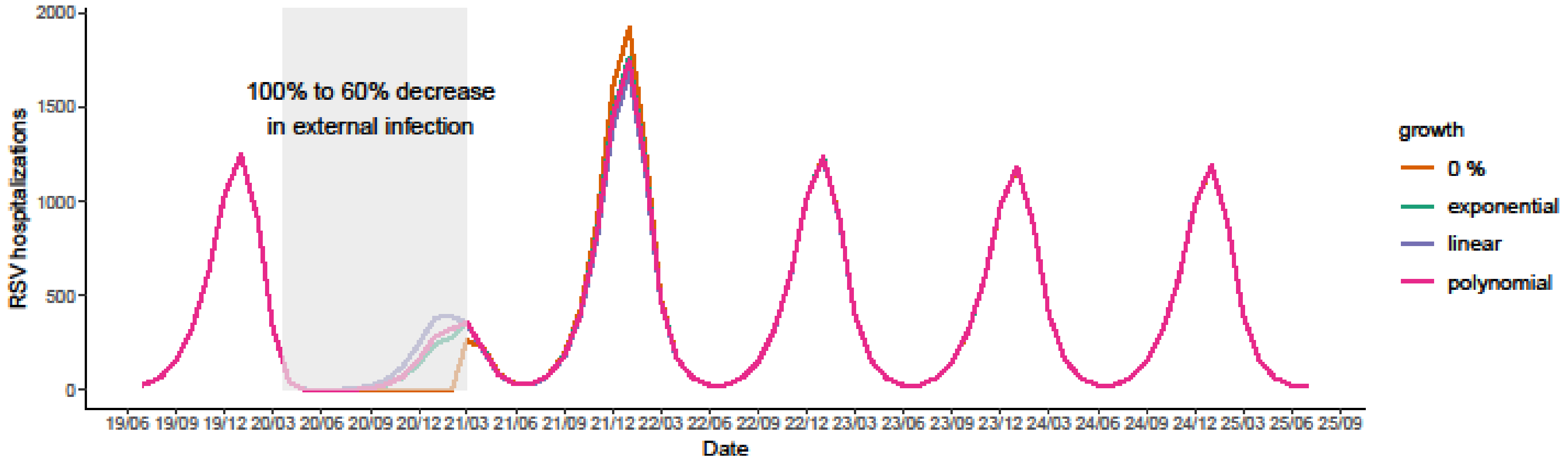


**Figure S6. External introduction of RSV infections on RSV epidemics.** This plot assumed a 5/100000 background external infections each month. The grey area indicates decreased external infections because of mitigation measures. The orange line shows the RSV epidemics if there were not external infections during April 1, 2020 to March 1, 2021. The green line, purple line and pink line indicate a sudden decrease in external infections at the beginning and a gradual increase with different growth rates.

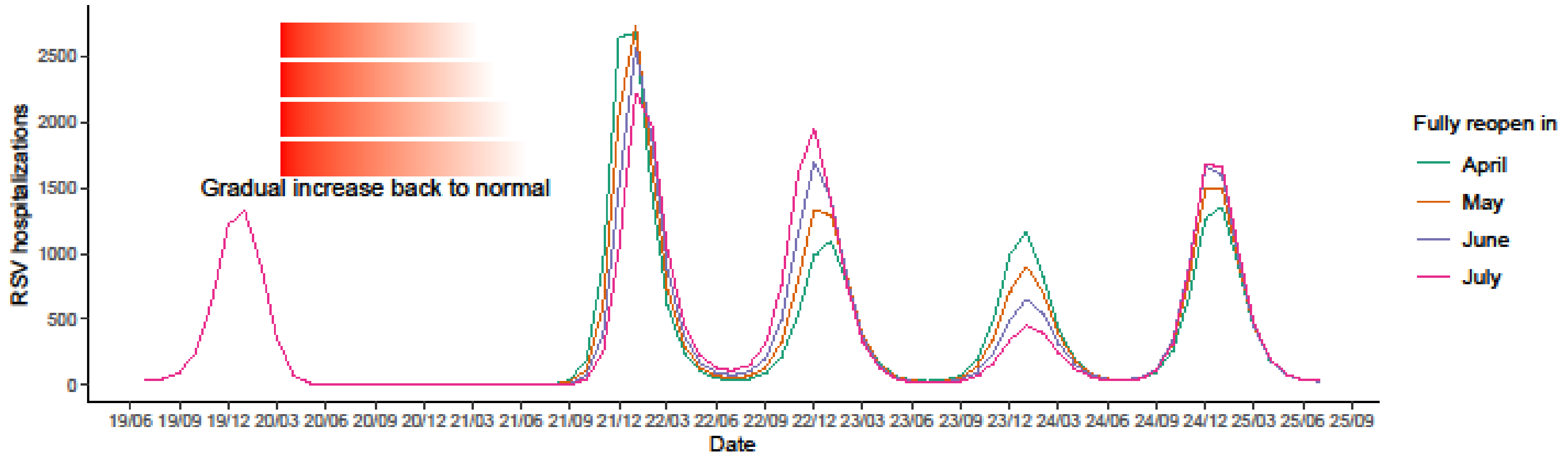


**Figure S7. The length of mitigation measures on RSV epidemics.** The colors of the lines show the expected RSV epidemics with different reopening dates in 2021 (first day of the month).

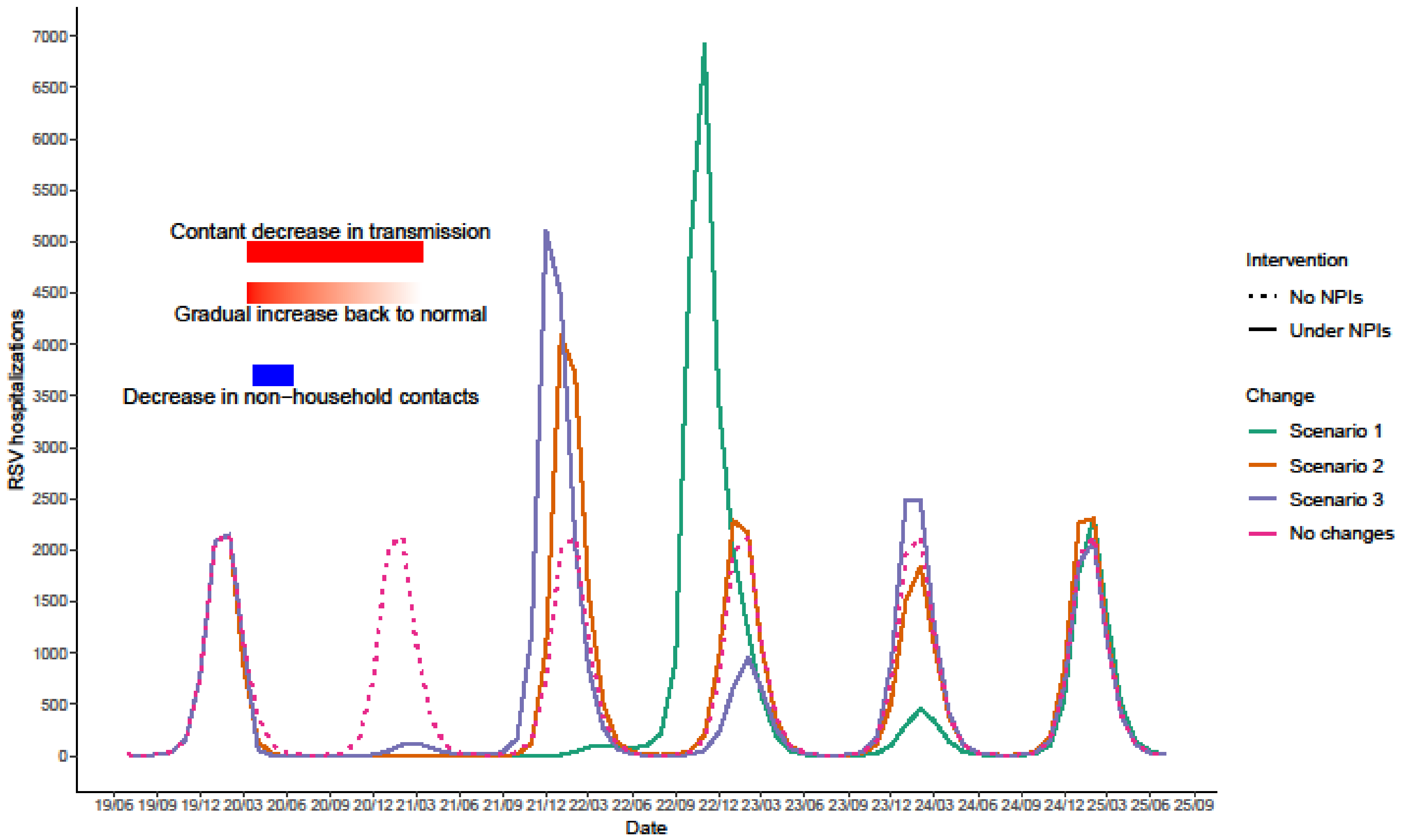


**Figure S8. Expected RSV hospitalizations under different scenarios, California, 2019–2025.** The dotted dark pink line shows the counterfactual scenario that there is no COVID-19 pandemic and no mitigation measures in place. The solid lines show three scenarios of stringency of mitigation measures. The green line represents Scenario 1: 20% constant decreased transmission from March 2020 to March 2021. The orange line represents Scenario 2: a sudden 20% decrease in RSV transmission in March 2020 followed by a linear increase back to normal. The purple line represents Scenario 3: 82% decreased non-household contacts and 10% increased household contacts between April and July 2020. The red rectangle on the top, the gradient red rectangle in the middle and the blue rectangle on the bottom indicate the length and the stringency of Scenario 1, Scenario 2 and Scenario 3, respectively.

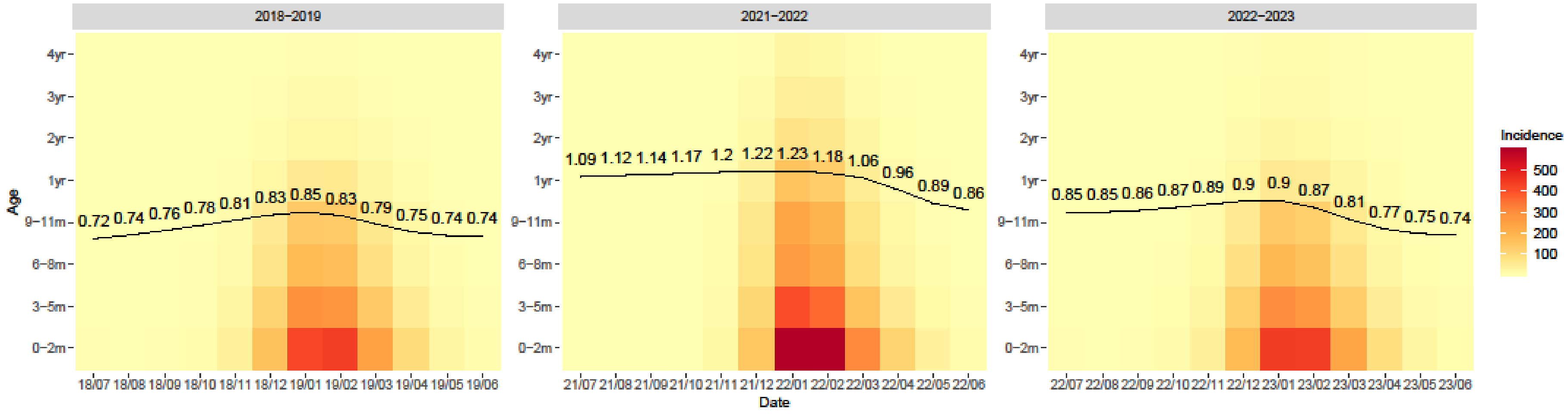


**Figure S9. The average age of hospitalization among children under 5 under Scenario 2, California.** The background color represents the incidence of RSV hospitalization in each age group in each month. The darker color suggests a higher incidence. The black line and value indicate the average age of hospitalization varies with time.

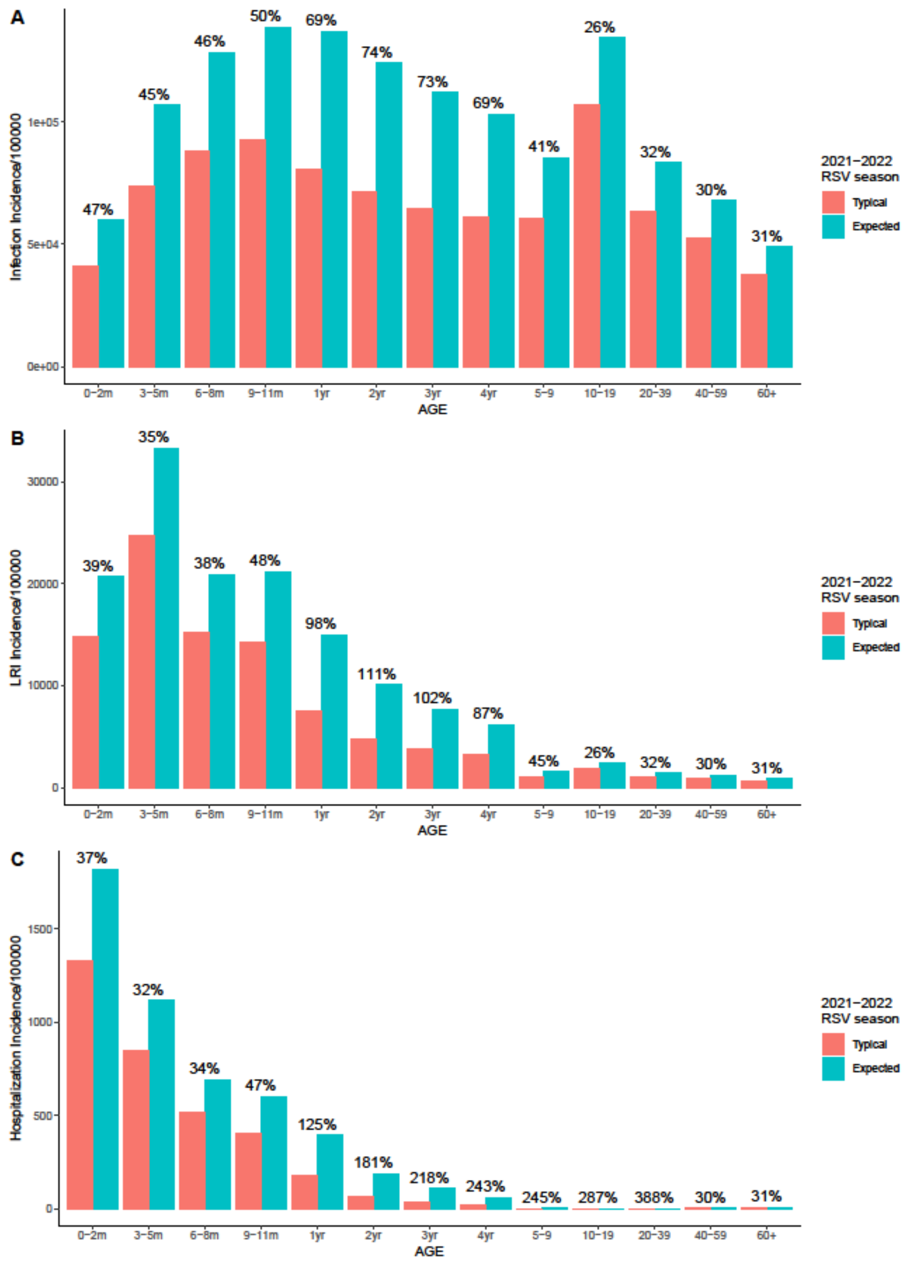
A
Infection Incidence/100000
AGE
2021−2022 RSV season
Typical
Expected
B
LRI Incidence/100000
C
Hospitalization Incidence/100000

**Figure S10. Age distribution of RSV infections, LRIs and hospitalizations, California, 2021-2022 RSV season.** Panel A to C correspond to RSV infections, RSV LRIs and RSV hospitalizations. The red color bars show the counterfactual incidence of RSV cases during 2021 to 2022 RSV season if there was no COVID-19 pandemic and no mitigation measures in place. The blue color bars show the expected incidence of RSV cases under Scenario 2 during 2021 to 2022 RSV season. The numbers on the top show the percentage difference between the expected incidence and the counterfactual incidence in each age group.

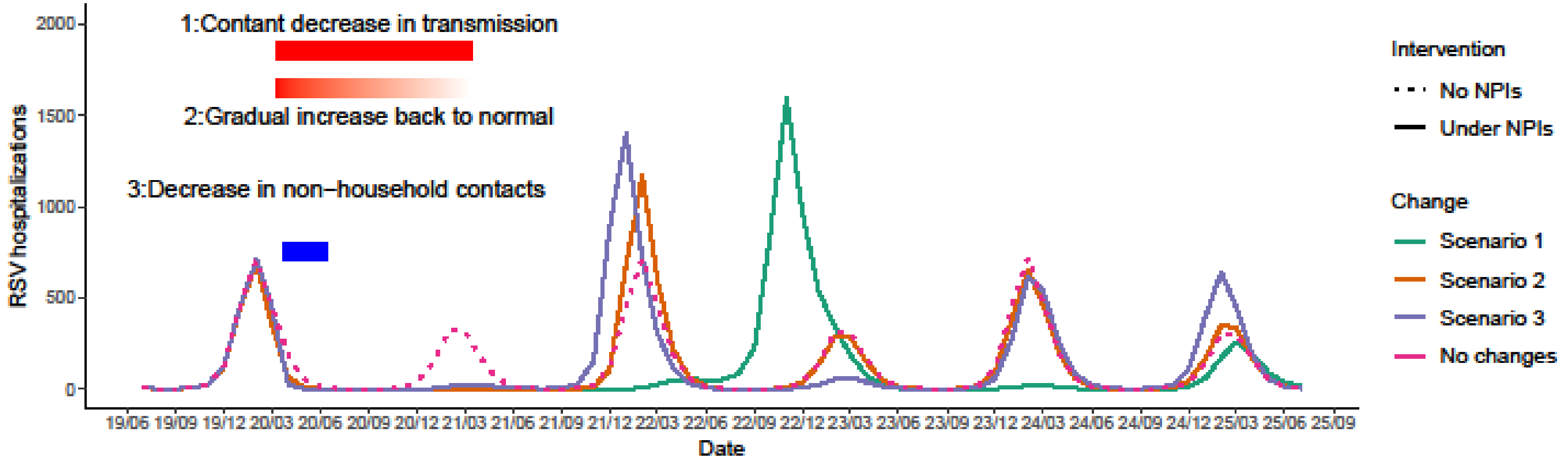


**Figure S11. Expected RSV hospitalizations under different scenarios for biennial epidemics peaking in even years, 2019–2025.** The dotted dark pink line shows the counterfactual scenario that there is no COVID-19 pandemic and no mitigation measures in place. The solid lines show three scenarios of stringency of mitigation measures. The green line represents Scenario 1: 20% constant decreased transmission from March 2020 to March 2021. The orange line represents Scenario 2: a sudden 20% decrease in RSV transmission in March 2020 followed by a linear increase back to normal. The purple line represents Scenario 3: 82% decreased non-household contacts and 10% increased household contacts between April and July 2020. The red rectangle on the top, the gradient red rectangle in the middle and the blue rectangle on the bottom indicate the length and the stringency of Scenario 1, Scenario 2 and Scenario 3, respectively.

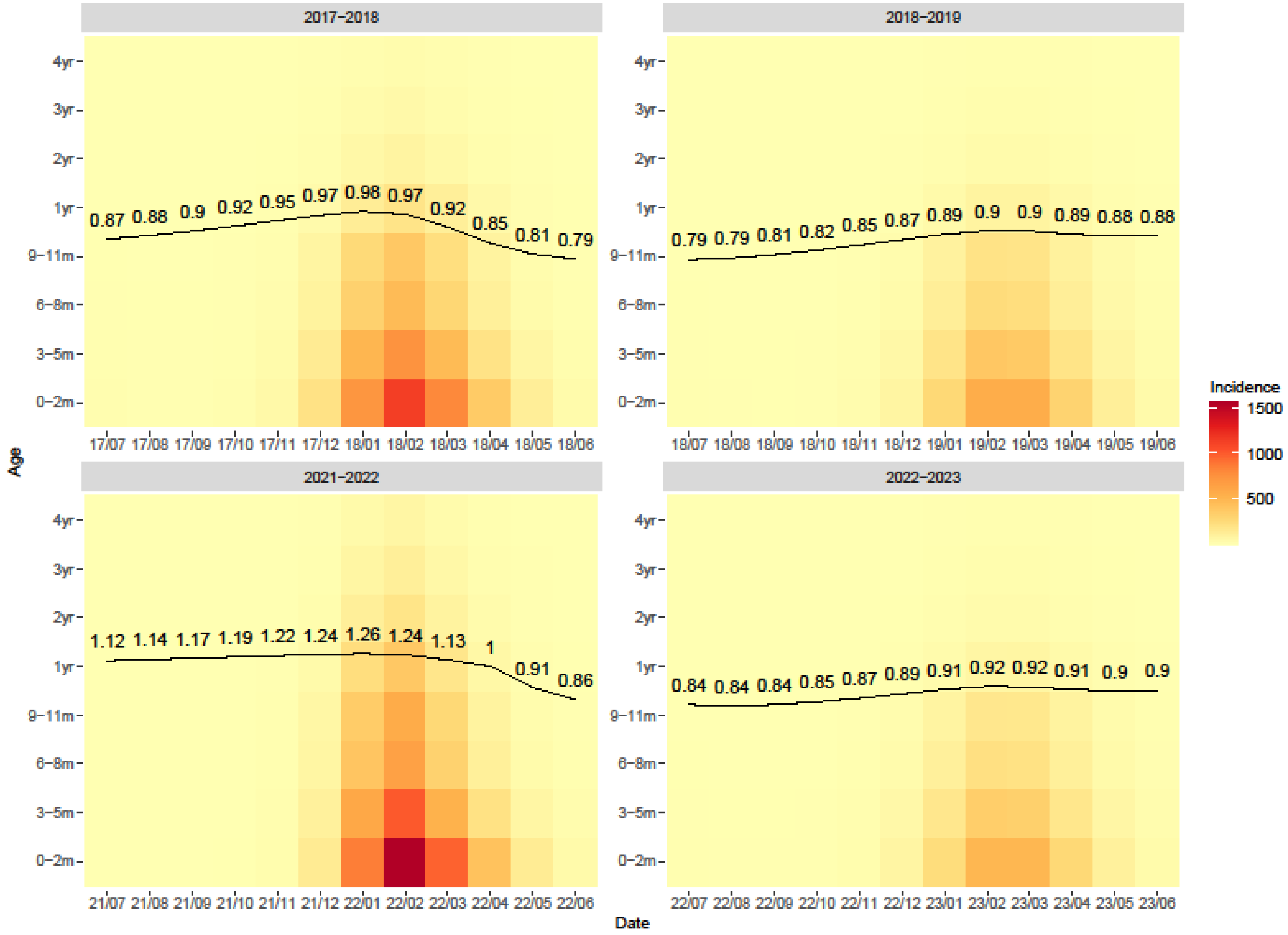


**Figure S12. The average age of RSV hospitalization for biennial epidemics peaking in even years among children under 5.** The background color represents the incidence of RSV hospitalization in each age group in each month. The darker color suggests a higher incidence. The black line and value indicate the average age of hospitalization varies with time.

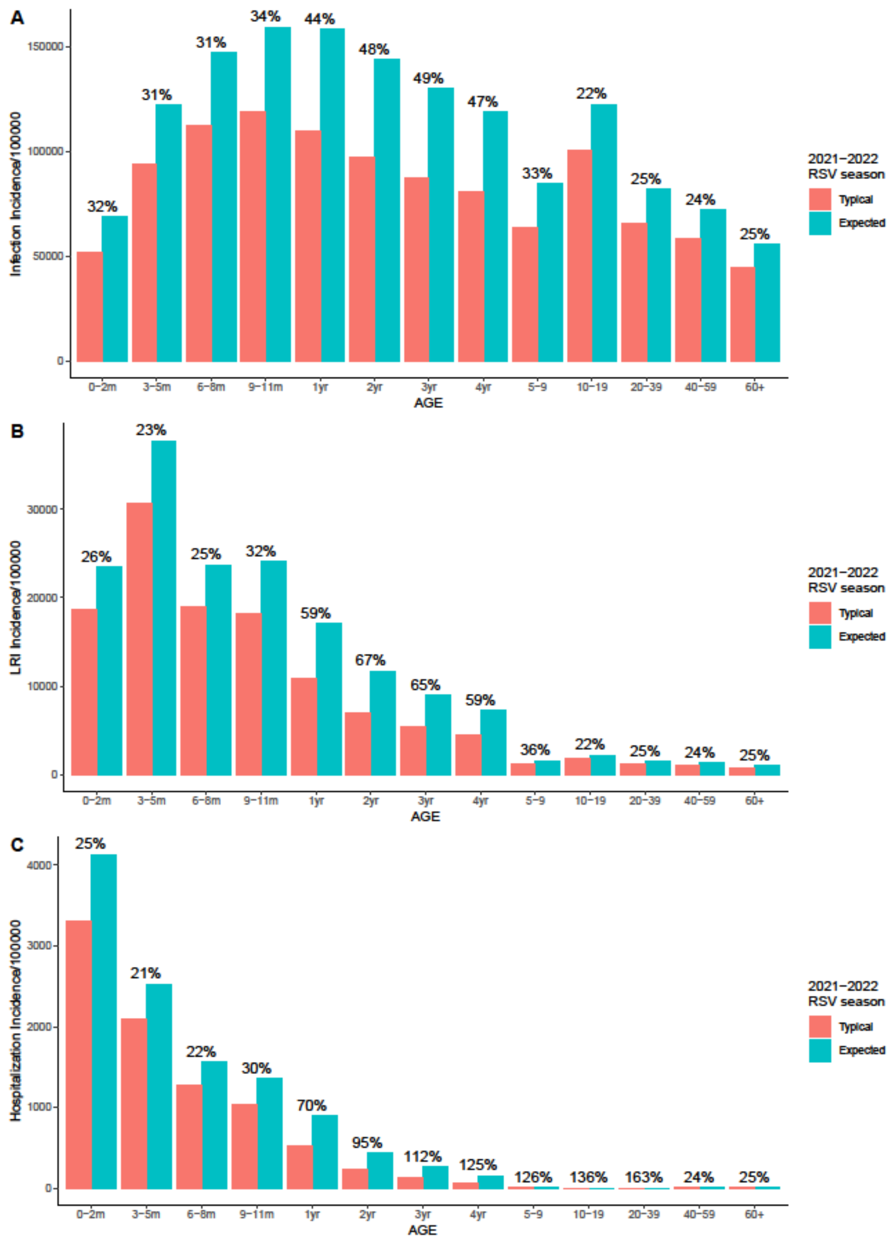
A
Infection Incidence/100000
150000
100000
50000
0
32%
31%
31%
34%
44%
48%
49%
47%
33%
22%
25%
24%
25%
0−2m
3−5m
6−8m
9−11m
1yr
2yr
3yr
4yr
5−9
10−19
20−39
40−59
60+
AGE
2021−2022
RSV season
Typical
Expected
B
LRI Incidence/100000
30000
20000
10000
0
26%
23%
25%
32%
59%
67%
65%
59%
36%
22%
25%
24%
25%
0−2m
3−5m
6−8m
9−11m
1yr
2yr
3yr
4yr
5−9
10−19
20−39
40−59
60+
AGE
2021−2022
RSV season
Typical
Expected
C
Hospitalization Incidence/100000
4000
3000
2000
1000
0
25%
21%
22%
30%
70%
95%
112%
125%
126%
136%
163%
24%
25%
0−2m
3−5m
6−8m
9−11m
1yr
2yr
3yr
4yr
5−9
10−19
20−39
40−59
60+
AGE
2021−2022
RSV season
Typical
Expected

114

**Figure S13. Age distribution of RSV infections, LRIs and hospitalizations under the assumption that biennial epidemics are greater in even years, 2021-2022 RSV season.** Panel A to C correspond to RSV infections, RSV LRIs and RSV hospitalizations. The red color bars show the counterfactual incidence of RSV cases during 2021 to 2022 RSV season if there was no COVID-19 pandemic and no mitigation measures in place. The blue color bars show the expected incidence of RSV cases under Scenario 2 during 2021 to 2022 RSV season. The numbers on the top show the percentage difference between the expected incidence and the counterfactual incidence in each age group.

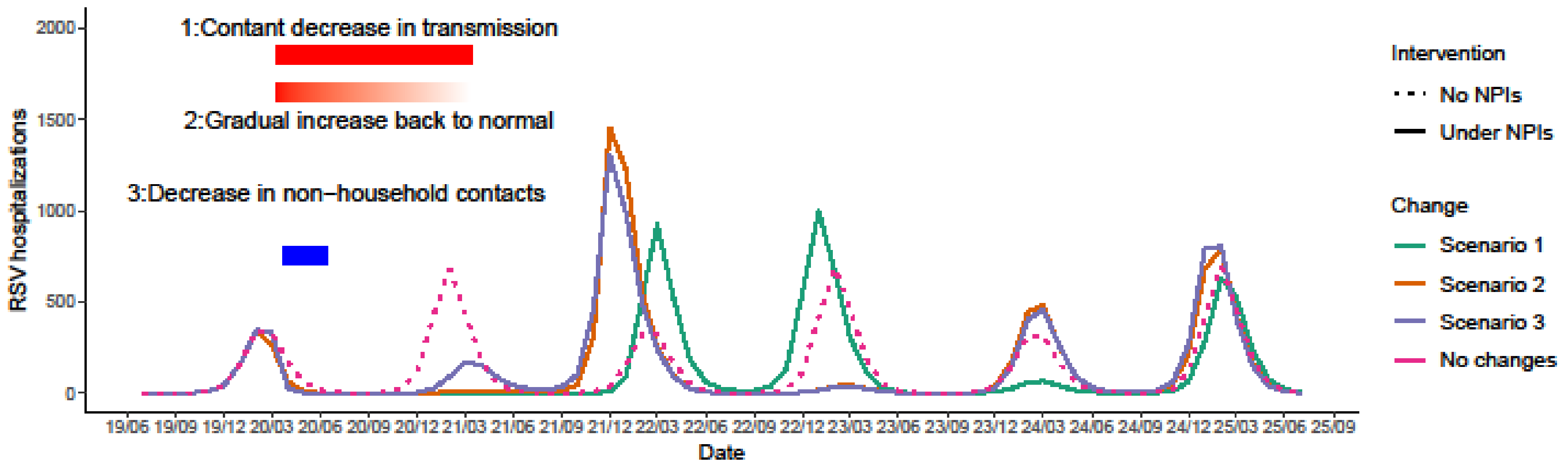


**Figure S14. Expected RSV hospitalizations under different scenarios for biennial epidemics peaking in odd years, 2019–2025.** The dotted dark pink line shows the counterfactual scenario that there is no COVID-19 pandemic and no mitigation measures in place. The solid lines show three scenarios of stringency of mitigation measures. The green line represents Scenario 1: 20% constant decreased transmission from March 2020 to March 2021. The orange line represents Scenario 2: a sudden 20% decrease in RSV transmission in March 2020 followed by a linear increase back to normal. The purple line represents Scenario 3: 82% decreased non-household contacts and 10% increased household contacts between April and July 2020. The red rectangle on the top, the gradient red rectangle in the middle and the blue rectangle on the bottom indicate the length and the stringency of Scenario 1, Scenario 2 and Scenario 3, respectively.

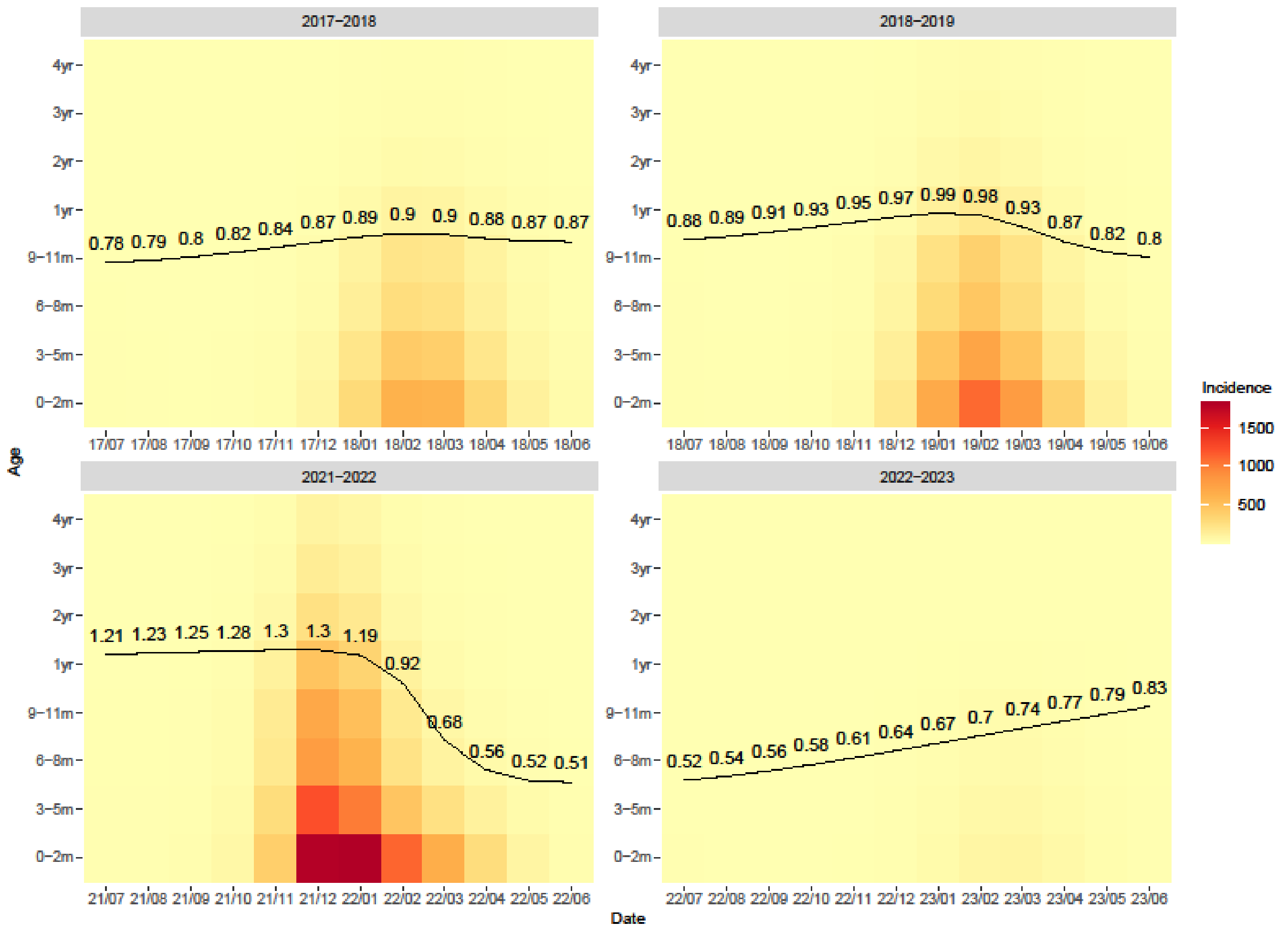


**Figure S15. The average age of RSV hospitalization for biennial epidemics peaking in odd years among children under 5.** The background color represents the incidence of RSV hospitalization in each age group in each month. The darker color suggests a higher incidence. The black line and value indicate the average age of hospitalization varies with time.

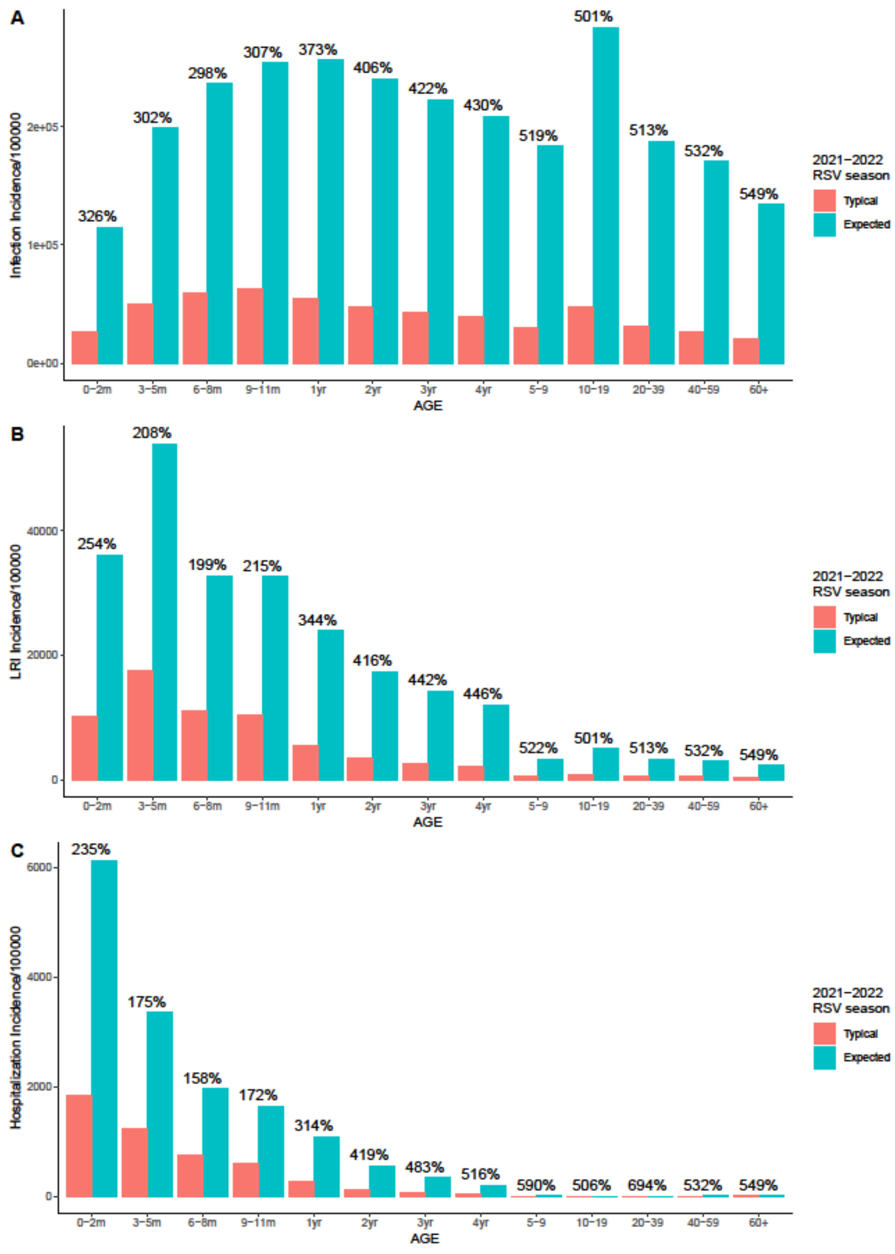

A
Infection Incidence/100000
0e+00
1e+05
2e+05
326%
302%
298%
307%
373%
406%
422%
430%
519%
501%
513%
532%
549%
0–2m
3–5m
6–8m
9–11m
1yr
2yr
3yr
4yr
5–9
10–19
20–39
40–59
60+
AGE
2021–2022
RSV season
Typical
Expected
B
LRI Incidence/100000
0
20000
40000
254%
208%
199%
215%
344%
416%
442%
446%
522%
501%
513%
532%
549%
0–2m
3–5m
6–8m
9–11m
1yr
2yr
3yr
4yr
5–9
10–19
20–39
40–59
60+
AGE
2021–2022
RSV season
Typical
Expected
C
Hospitalization Incidence/100000
0
2000
4000
6000
235%
175%
158%
172%
314%
419%
483%
516%
590%
506%
694%
532%
549%
0–2m
3–5m
6–8m
9–11m
1yr
2yr
3yr
4yr
5–9
10–19
20–39
40–59
60+
AGE
2021–2022
RSV season
Typical
Expected

**Figure S16. Age distribution of RSV infections, LRIs and hospitalizations under the assumption that biennial epidemics are greater in odd years, 2021-2022 RSV season.** Panel A to C correspond to RSV infections, RSV LRIs and RSV hospitalizations. The red color bars show the counterfactual incidence of RSV cases during 2021 to 2022 RSV season if there was no COVID-19 pandemic and no mitigation measures in place. The blue color bars show the expected incidence of RSV cases under Scenario 2 during 2021 to 2022 RSV season. The numbers on the top show the percentage difference between the expected incidence and the counterfactual incidence in each age group.
